# Development and Evaluation of a Regional Health Information Exchange (HIE)-Based Platform for Population-Level Infectious Disease Surveillance

**DOI:** 10.64898/2026.10.04.26364717

**Authors:** Micaela Sandoval, Jenny Mikhail, Hao He, Courtney Denton, Haider al Rustem, Tru Cao, Catherine Troisi, Jennifer Deegan, Blake Hanson, Cici Bauer, Eric Boerwinkle

## Abstract

**Background:** Health Information Exchanges (HIEs) offer a potential resource for timely, population-level infectious disease surveillance by integrating electronic health record data across healthcare systems.

**Methods:** We developed an end-to-end informatics pipeline for near-real-time community-level health surveillance based in a regional HIE serving Southeast Texas. We identified deidentified individual-level COVID-19, influenza, and RSV cases using diagnosis codes and laboratory results and assessed demographic and temporal trends from January 2022 through August 2026.

**Results:** We identified 492,242 COVID-19 cases, 343,977 influenza cases, and 33,159 RSV cases during the study period. Surveillance data captured expected seasonal patterns, including recurring winter COVID-19 and influenza peaks and shifting RSV epidemiology, while enabling monitoring of laboratory test positivity metrics and hospitalization trends across age groups.

**Conclusions:** HIE-based surveillance provides scalable, near-real-time monitoring of disease targets across large populations. HIE infrastructure can enhance public health situational awareness and complement traditional surveillance systems by harmonizing vital health data across systems and providers and delivering timely, standardized, and representative population health indicators.

## Introduction

Timely surveillance is essential for identifying emerging infectious disease threats, monitoring disease trends, and informing public health response. Public health agencies rely on multiple surveillance systems, including passive reporting of notifiable conditions, laboratory reporting, syndromic surveillance, and environmental surveillance such as wastewater monitoring. However, these systems vary in timeliness, population coverage, and the availability of individual-level clinical information. Electronic health record (EHR) datasets can potentially bridge surveillance gaps by providing near-real-time, individual-level clinical and laboratory data that can be translated into surveillance indicators for a wide range of health conditions [1–5].

The use of EHR data for public health applications also presents several challenges. EHR datasets are typically limited to a single electronic medical record (EMR) or healthcare system, with information loss occurring as patients move between healthcare systems [6,7]. Additionally, accurate analysis of high-dimensional EHR data requires significant processing and data science capacity, creating barriers for public health agencies without the necessary infrastructure. Health information exchanges (HIEs) are local, regional, state, or national networks that ingest, harmonize, and exchange clinical data across multiple healthcare systems. While the primary objective of any HIE is care coordination, these large EHR datasets are increasingly being utilized for population-based health services, epidemiological, and operational research [8–12].

In this study, we describe the development and implementation of a near-real-time infectious disease surveillance platform utilizing data from a regional HIE in Southeast Texas. We describe the development of the underlying data infrastructure, assess HIE population coverage, and examine trends in COVID-19, influenza, and respiratory syncytial virus (RSV) from 2022 through 2026.

## Methods

In collaboration with Healthconnect Texas (HTX), the regional HIE for Southeast Texas, we developed a flexible, sustainable, and comprehensive platform for near-real-time analysis of EHR-based health trends. UTHealth Houston School of Public Health initially partnered with HTX, the regional health information exchange (HIE) for Southeast Texas to develop a longitudinal, individual-level, query-able cohort dataset anchored to a master patient index (MPI) containing >25 million unique patients to facilitate population-based COVID-19 clinical epidemiology studies [13,14]. Recognizing the potential of this large, representative, regionally-specific clinical database for public health surveillance, the UTHealth Houston SPH and HTX teams collaborated to create a tailored, near-real-time surveillance platform for population-based epidemiology, health services research, and public health decision-support. The objective was to create a foundation of prescreened EHR data sources which could be accessed through a basic query structure and formatted for a wide variety of epidemiological and population health studies. To pilot this novel health informatics platform, we investigated population-level trends in respiratory infections, specifically COVID-19, influenza, and respiratory syncytial virus (RSV) disease in the Greater Houston Area for the period from January 2022 through August 2026.

### Preliminary assessment of population-level EHR data

#### Population coverage assessment

To support population-based studies in the Greater Houston Area, we first assessed the patient population covered by the regional HIE, HTX. For this investigation, the study area was Harris County, Texas and the surrounding counties, including Fort Bend, Galveston, Montgomery, Liberty, Waller, Chambers, and Brazoria counties, home to more than 7 million residents including the city of Houston. Coverage was assessed among all unique individuals with at least one clinical encounter recorded in HTX during 2024.

Patients with a qualifying 2024 encounter were aggregated by census tract of residence, sex, and age as of July 1, 2024, represented categorically as 5-year bands. Sixteen of the 1,594 tracts with no residential population, predominantly group-quarters populations, or non-residential land use were excluded. Patient demographics were compared against both the 2020 Decennial Census counts [15] and 2019–2023 American Community Survey (ACS) 5-year estimates [16–17]. For this assessment, coverage was defined within each stratum as patients with a qualifying encounter over either the Census or ACS population estimate, expressed as a percentage. Tract-level representativeness was additionally calculated by comparing the HTX patient population against the ACS 90% confidence interval [18]. Individuals with unknown sex were included in overall and age-specific coverage but excluded from age–sex comparisons.

#### Evaluation of feature integrity and target selection

EHR data is complex, made up of many distinct features, such as diagnoses, laboratory feeds, encounter information, procedures, imaging studies, etc. However, the availability of specific data streams within the HIE varies widely across contributing healthcare providers. We therefore conducted an *a priori* investigation of the availability, integrity, and interpretability of specific features, compared to expected results from literature or population health studies. We utilized an age-stratified, random sample of Harris County residents captured by the HIE (n=700,000 unique patients) to examine the proportion of patients and events, such as hospital admissions, with associated diagnosis, encounter, laboratory test result, procedure, medication, vital signs, social history, imaging, vaccination history, and clinical notes recorded in the longitudinal record. We determined that features contained within minimum data sets such as Admission/Discharge/Transfer (ADT) feeds, including diagnoses, encounter data, and patient demographics were typically the most robust and consistently updated, while text- or image-based features such as imaging or clinical notes were less available and may not be suitable for population-based work. Some features, including laboratory test results, were not consistently available for all patient encounters, limiting their utility for individual-level analyses, e.g., to determine that a specific patient did not test positive for a pathogen during a hospitalization.

However, we did explore the potential utility of population-based metrics derived from these features, including laboratory test positivity rates, given the large sample size of test results extracted from the Consolidated Continuity of Care Documents (C-CDA).

We utilized an age-stratified, random sample of Harris County residents captured by the HIE to 1) explore coverage and integrity of various features and 2) develop and validate case detection algorithms (CDAs). Finally, we implemented an end-to-end informatics pipeline for near-real-time querying, deidentification, and processing of respiratory infection diagnoses, as well as laboratory test data for selected pathogens.

### Target extraction

#### Case detection algorithms

The following respiratory infectious disease targets were selected based on the feature evaluation described above: 1. diagnoses of COVID-19, influenza, or RSV; and 2. diagnostic laboratory test results for SARS-CoV-2, Influenza A, Influenza B, or RSV. Case detection algorithms were developed and validated in an age-stratified, random sample of Harris County residents captured by the HIE. COVID-19, influenza, and RSV cases were defined as any patient with a diagnosis identified through ICD-10 or SNOMED-CT codes. For this investigation, only diagnoses associated with acute infection were included, i.e., *U07*.*1: COVID-19, virus identified* would be included but *U08*.*9: Personal history of COVID-19, unspecified* would not be included.

Diagnostic antigen and nucleic acid amplification laboratory tests were identified though LOINC © or SNOMED-CT codes; antibody tests were excluded in this analysis.

#### Target extraction and deidentification

All targets were extracted via weekly queries to the live HIE database. The HTX team executed these queries and created deidentified, line-listing-style, individual-level datasets containing unique patient IDs, diagnosis codes and text descriptions, laboratory test orders and results, encounter types (inpatient, outpatient, or emergency), and patient demographics including age, sex, and census tract. Data was pulled weekly, using the *Morbidity and Mortality Weekly Report* (MMWR) weeks [19], and no exact dates or other identifying information was included. The UTHealth team received these deidentified datasets through a secure file transfer protocol site and implemented a custom informatics pipeline for processing, visualization, and analysis.

#### De-duplication and episode analysis

Diagnosis-level records were de-duplicated by unique patient ID and MMWR week to generate case counts stratified by age, sex, and geography. Encounter types were reduced into non-exclusionary categories, i.e., counts of patients with any COVID-19 diagnosis recorded at an inpatient encounter, to accommodate individuals with multiple related encounters in a week.

Individual-level COVID-19, influenza, and RSV records were additionally condensed into 2-week episodes to reduce duplication from individuals with multiple related encounters across two reporting periods.

Laboratory test positivity percentage was calculated as the total number of unique individuals with at least one qualifying positive test divided by the total number of unique individuals with at least one qualifying test reported within the reporting period. Positivity rates for each pathogen (SARS-CoV-2, RSV, Influenza A and Influenza B) were stratified by patient age, geography, and assay type (antigen vs. nucleic acid amplification test).

### Study population

The study area was the Greater Houston Area, defined as Harris County, Texas and the seven surrounding counties. Census tract derived from a geocoded patient-reported residential address captured at the date of data pull was the primary geographic unit for this analysis. For patients whose addresses could not be geocoded, ZIP code roughly matched to county boundaries was used as a proxy to determine eligibility. Individuals were included in this analysis if their self-reported residential address was within the catchment area and they had at least one target, either diagnosis or laboratory test report, recorded in the HIE between January 1, 2022 and August 26, 2026.

### Statistical analysis

Demographic and clinical data were reported as frequencies and proportions for categorical variables. Case count time series for COVID-19, influenza, and RSV are presented as 4-week moving averages stratified by demographics. Laboratory test positivity rates and total test count time series are presented as 4-week moving averages. Geospatial analyses were conducted utilizing individual, de-duplicated patients’ residential addresses. Census data were retrieved using the tidycensus R package [20]. Small cell values (1 to 4 individuals per cell) were suppressed to preserve patient privacy. All statistical analyses were performed using Python and RStudio.

### Ethics statement

This study using deidentified EHR data for infectious disease monitoring was approved by the University of Texas Health Science Center at Houston Committee for Protection of Human Subjects as a public health project and granted a waiver of informed consent (Study number: HSC-IMM-24-0381).

## Results

### Population coverage assessment

Within the Greater Houston study area, which included Harris, Fort Bend, Galveston, Montgomery, Liberty, Waller, Chambers, and Brazoria counties, the HTX network demonstrated high coverage of hospitals, including rural and urban health centers, hospital-based emergency departments, private health systems, and county hospitals. Over 85% of non-psychiatric inpatient beds assessed by the Texas Department of State Health Services in these counties were within HTX participating institutions in 2025. Additionally, the HTX network included over 2,000 ambulatory practices including standalone emergency departments and urgent care centers, primary care practices, and specialty practices.

In the 1,578 analyzed census tracts, 4,129,143 unique individuals had at least one clinical encounter captured in the HIE in 2024, corresponding to 58.6% of the 2020 Census population and 57.6% of the 2019–2023 ACS estimate. Coverage was lowest among school-aged children and adolescents aged 5–19 years (40.3%) and highest among adults aged 65 and older (91.1%). Women were more heavily represented compared to men from adolescence onward; this discrepancy was widest at ages 20–39 (women 66.8% vs men 40.4% coverage) and closed by age 80. Coverage patterns by age and sex were nearly identical against ACS estimates. Median tract-level coverage was 56.2% (IQR 49.5–63.6); 2.9% of tracts had coverage below 40% and 1.5% at or above 100%.

### Respiratory infection case detection and laboratory test positivity

Our novel HIE-based surveillance platform identified 492,242 unique individuals diagnosed with COVID-19, 343,977 diagnosed with influenza, and 33,159 diagnosed with RSV in the Greater Houston Area between January 1, 2022 and August 26, 2026. Yearly case counts stratified by pathogen, demographics, and clinical features are presented along with laboratory test result summaries in Tables 1-3. Temporal trends in COVID-19, influenza, and RSV episodes by year are presented in Figure 2. Age-stratified trends in counts of inpatient hospitalization episodes for each pathogen are presented in Figure 3. Case counts derived from diagnoses are displayed alongside counts of total patients with at least one antigen or PCR test resulted and test positivity percentage in Figure 4.

**Table 1:**
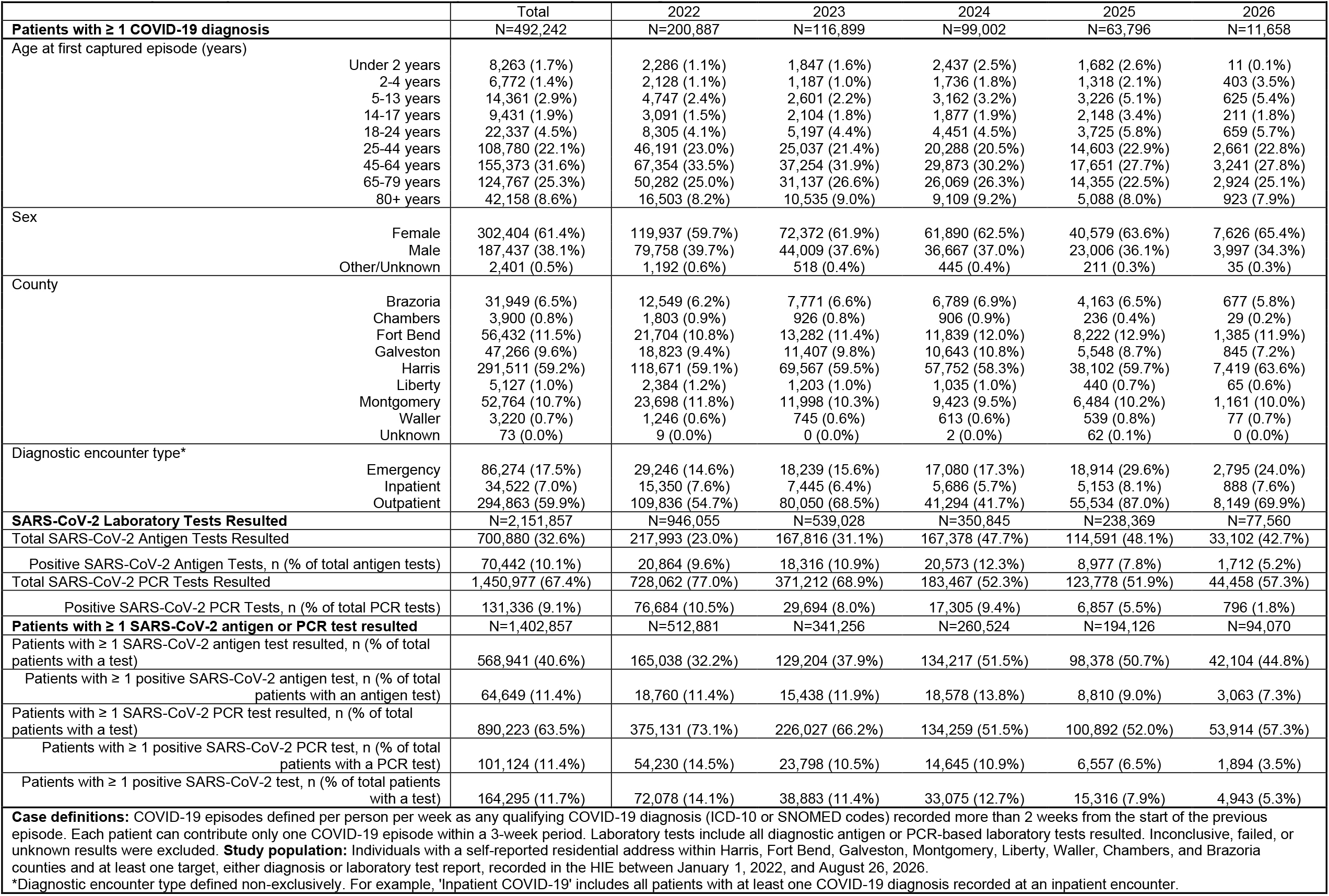
COVID-19 episodes captured in HIE-based surveillance platform, January 2022 to August 2026.

**Figure 1.**
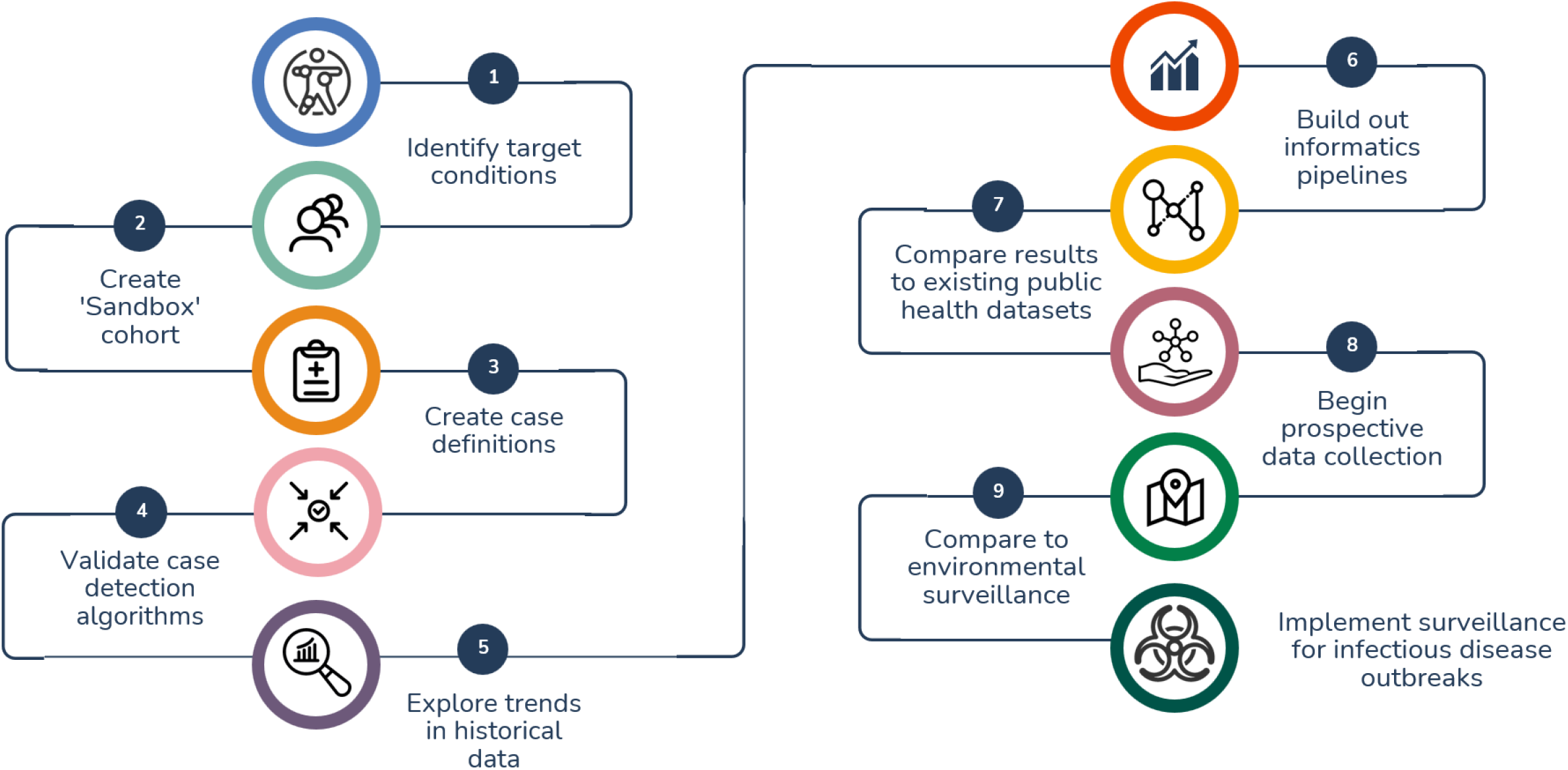
Flow chart depicting development of near-real-time infectious disease surveillance platform from a health information exchange

**Figure 2.**
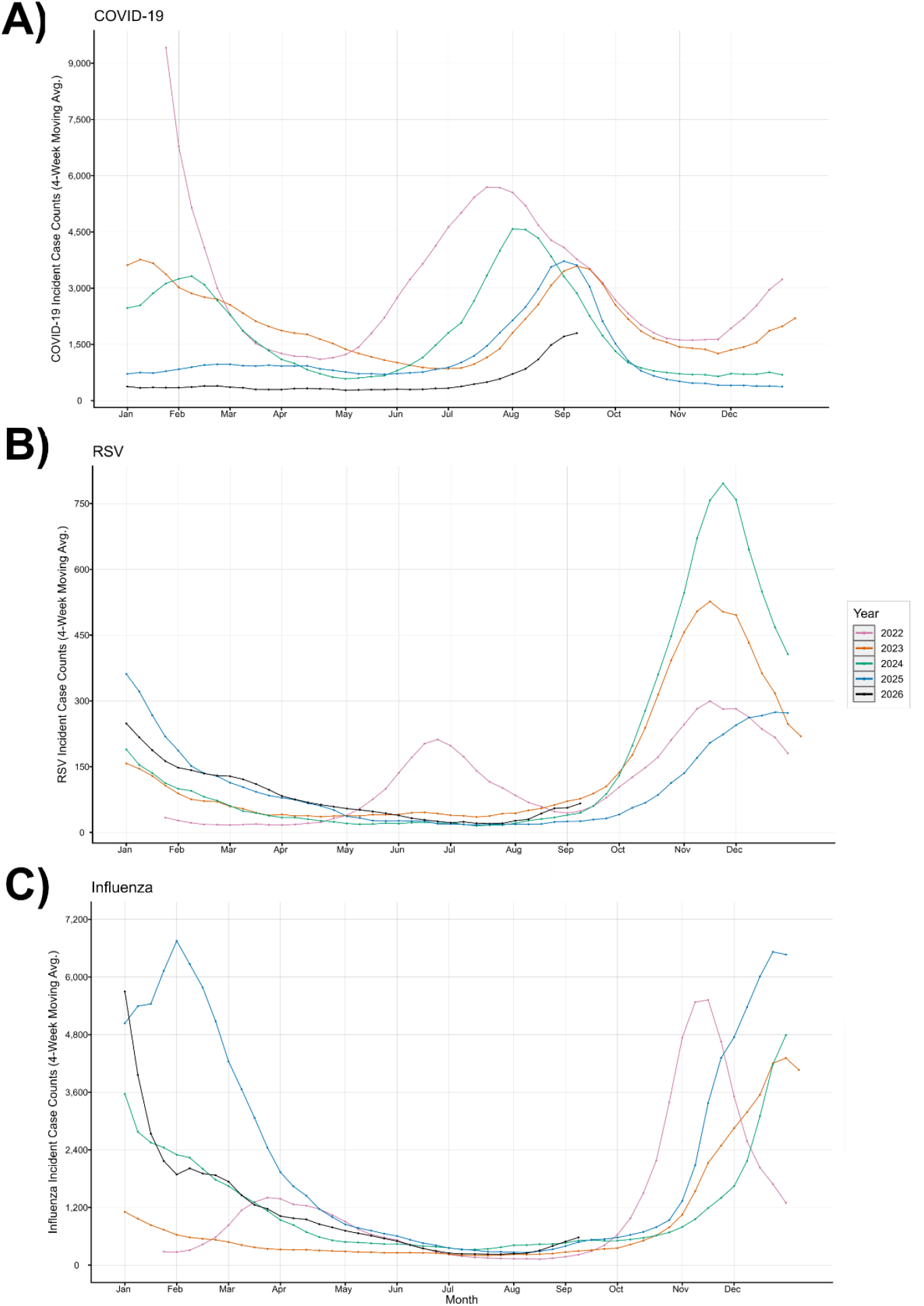
COVID-19, influenza, and RSV cases: 4 week moving average of incident cases, Greater Houston January 2022-August 2026. Caption: Episodes defined per person per week as any qualifying COVID-19, RSV or influenza diagnosis (ICD-10 or SNOMED codes) recorded more than 2 weeks from the start of the previous episode. Each patient can contribute only one episode within a 3-week period. Includes individuals with a self-reported residential address within Harris, Fort Bend, Galveston, Montgomery, Liberty, Waller, Chambers, and Brazoria counties and at least one COVID-19 diagnosis, recorded in the HIE between January 1, 2022 and August 26, 2026.

**Figure 3.**
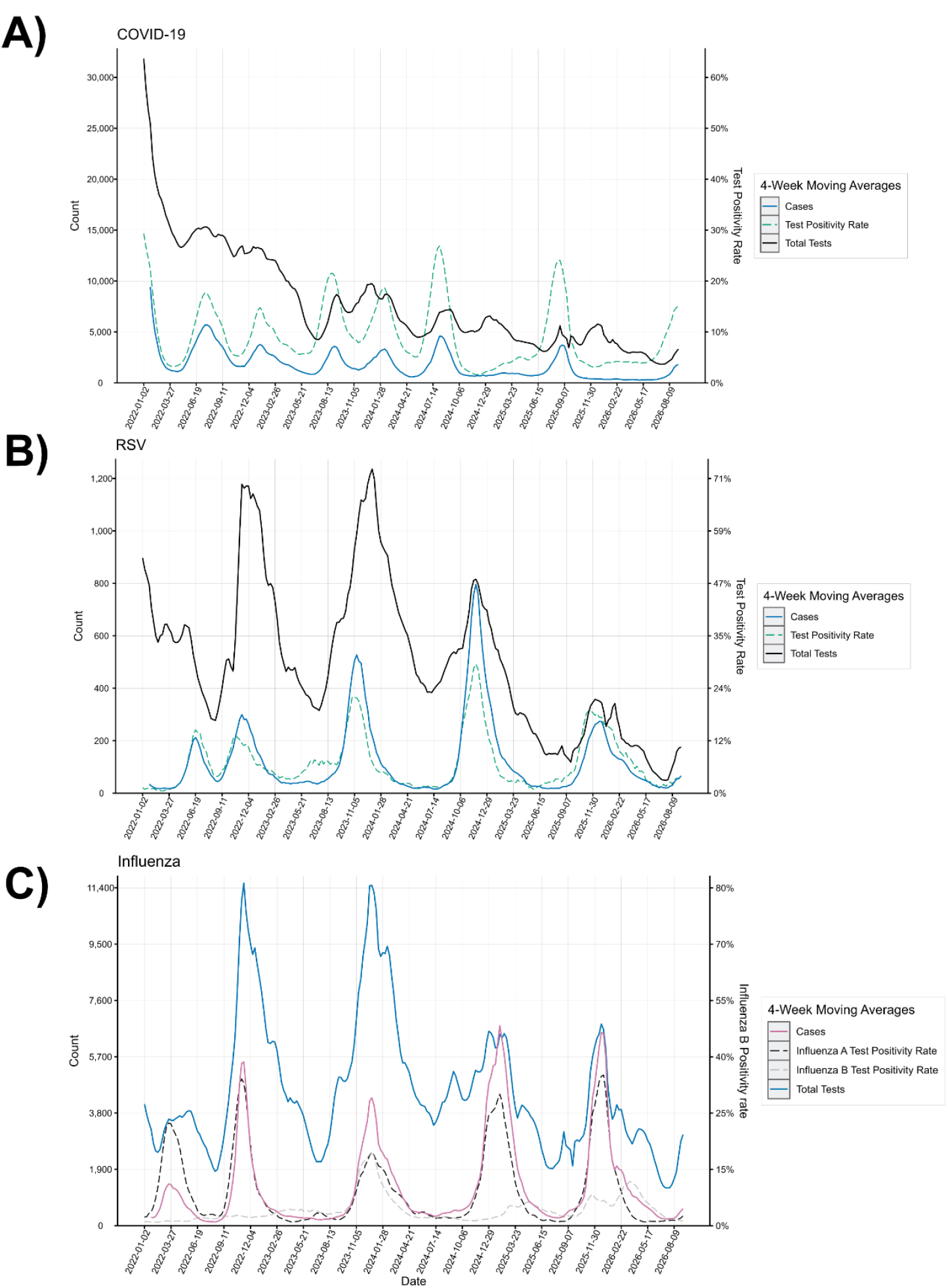
COVID-19, RSV, and influenza cases: 4 week moving average of incident cases, Greater Houston January 2022-August 2026. Caption: Episodes defined per person per week as any qualifying COVID-19, RSV or influenza diagnosis (ICD-10 or SNOMED codes) recorded more than 2 weeks from the start of the previous episode. Laboratory tests include all diagnostic antigen or PCR-based laboratory tests resulted. Inconclusive, failed, or unknown results were excluded. Laboratory test positivity defined as # of individuals with ≥ 1 positive test/ # of individuals with ≥ 1 test resulted, calculated weekly. Includes individuals with a self-reported residential address within Harris, Fort Bend, Galveston, Montgomery, Liberty, Waller, Chambers, and Brazoria counties and at least one target, either diagnosis or laboratory test report, recorded in the HIE between January 1, 2022 and August 26, 2026.

**Figure 4.**
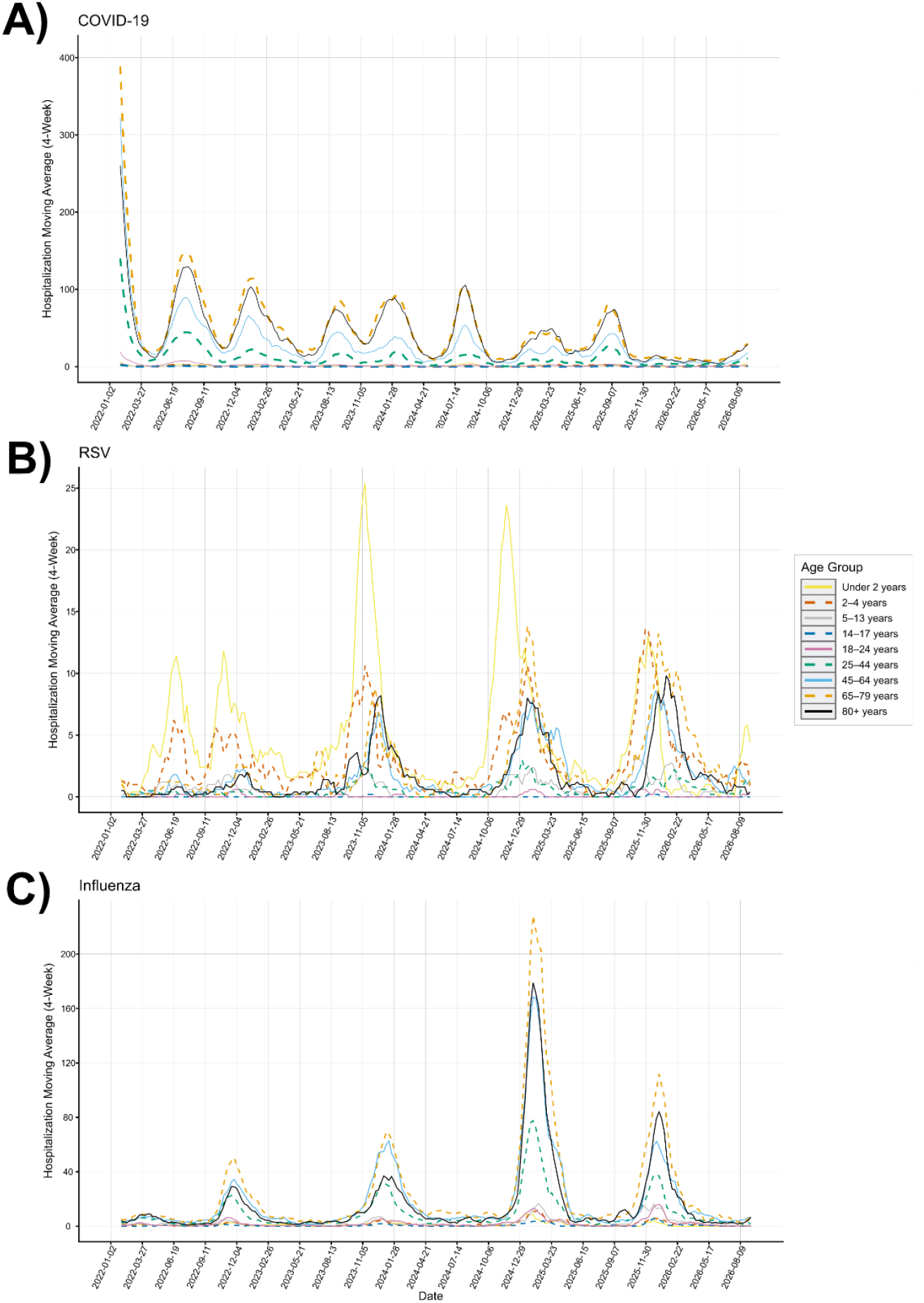
COVID-19, RSV, and influenza associated inpatient hospitalizations: 4 week moving average of incident cases stratified by age, Greater Houston January 2022-August 2026. Caption: Hospitalizations defined per person per week as any qualifying inpatient admission with an associated COVID-19, RSV or influenza diagnosis (ICD-10 or SNOMED codes) recorded more than 2 weeks from the start of the previous episode. Age stratification based on age at beginning of reporting week. Includes individuals with a self-reported residential address within Harris, Fort Bend, Galveston, Montgomery, Liberty, Waller, Chambers, and Brazoria counties and at least one target, either diagnosis or laboratory test report, recorded in the HIE between January 1, 2022 and August 26, 2026.

#### COVID-19

COVID-19 case counts were significantly higher in 2022 than in the later years dropping from 200,887 in 2022 to between 63,796 and 116,899 in 2023-2025 (Table 1). While the total case counts dropped dramatically, the age, sex, and geographic case distributions were relatively stable across time. COVID-19 cases were more likely to have at least one associated inpatient or emergency department encounter in 2025, compared with the other years. In total, 2,151,857 antigen or PCR diagnostic SARS-CoV-2 tests were recorded over the study period, with the proportion of antigen tests rising from 23% in 2022 to 48% in 2025. COVID-19 case peaks shifted substantially over the observation period (Figure 2). In January of 2022, there was a very high caseload, and there were noticeable winter peaks in January-February of 2023 and 2024.

However, there did not appear to be any significant COVID-19 peaks associated with the late winter of 2025 or 2026. In contrast, a summer peak appeared in all years, though the magnitude seemed to decrease over time; the summer case influx additionally shifted later over time, with the peak falling in July in 2022 and in mid-September in 2026. SARS-CoV-2 laboratory test positivity appears to follow case trends, though total test volume varied widely over the study period (Figure 3). COVID-19-associated inpatient encounters also followed case trends, with disproportionately high hospitalizations among people over 65 years and a marked decline from the spike in January 2022 (Figure 4).

#### RSV

RSV typically displays a strong seasonal trend, with cases peaking in the late fall/winter months. However, in our study population, there appears to be a lull in RSV cases during the COVID-19 pandemic, followed by an unusual summer peak and a relatively small fall peak in 2022 (Table 2). Children under 5 years of age represent the large majority of RSV cases in all years, although the proportion of RSV cases among infants under 2 years decreased from around 45% in 2022-2024 to 29% in 2025 and only 9% in 2026. Of note, the proportion of RSV cases with an associated inpatient hospitalization was higher in 2025 and 2026, compared with previous years. As with SARS-CoV-2, a greater proportion of antigen tests for RSV were reported each progressive year (n=165,456 total antigen and PCR tests). Following the atypical activity in summer, RSV cases peaked consistently in late fall for 2022-2025; the 2025 fall peak was demonstrably lower magnitude than in previous years (Figure 2). While the laboratory test positivity rate for RSV approximately followed case counts, the temporal alignment displayed more variability than in SARS-CoV-2 (Figure 3). RSV-associated hospitalizations exhibited disparate trends across epidemic peaks, with children under 5 appearing to lead the epidemic curve in most peak periods. Hospitalizations within the summer and fall 2022 peaks were almost exclusively among young children, and infants and children under 5 as well as adults over 65 were disproportionately represented in 2023, 2024, and 2025 (Figure 4). The number of RSV-associated hospitalizations among infants under 2 declined in 2025, though the hospitalization count among children 2-4 years old remained stable.

**Table 2:** RSV episodes captured in HIE-based surveillance platform, January 2022 to August 2026.

|  | Total | 2022 | 2023 | 2024 | 2025 | 2026 |
| --- | --- | --- | --- | --- | --- | --- |
| **Patients with ≥ 1 RSV diagnosis** | **N=33,159** | **N=6,293** | **N=8,307** | **N=9,986** | **N=5,741** | **N=2,832** |
| Age at first captured episode (years) |  |  |  |  |  |  |
| Under 2 years | 13,261 (40.0%) | 3,013 (47.9%) | 3,655 (44.0%) | 4,656 (46.6%) | 1,685 (29.4%) | 252 (8.9%) |
| 2-4 years | 10,510 (31.7%) | 1,937 (30.8%) | 2,363 (28.4%) | 2,986 (29.9%) | 2,065 (36.0%) | 1,159 (40.9%) |
| 5-13 years | 2,421 (7.3%) | 573 (9.1%) | 540 (6.5%) | 661 (6.6%) | 424 (7.4%) | 223 (7.9%) |
| 14-17 years | 164 (0.5%) | 23 (0.4%) | 37 (0.4%) | 47 (0.5%) | 36 (0.6%) | 21 (0.7%) |
| 18-24 years | 303 (0.9%) | 28 (0.4%) | 69 (0.8%) | 89 (0.9%) | 68 (1.2%) | 49 (1.7%) |
| 25-44 years | 1,192 (3.6%) | 136 (2.2%) | 323 (3.9%) | 295 (3.0%) | 245 (4.3%) | 193 (6.8%) |
| 45-64 years | 1,928 (5.8%) | 248 (3.9%) | 477 (5.7%) | 429 (4.3%) | 446 (7.8%) | 328 (11.6%) |
| 65-79 years | 2,195 (6.6%) | 224 (3.6%) | 538 (6.5%) | 530 (5.3%) | 534 (9.3%) | 369 (13.0%) |
| 80+ years | 1,185 (3.6%) | 111 (1.8%) | 305 (3.7%) | 293 (2.9%) | 238 (4.1%) | 238 (8.4%) |
| Sex |  |  |  |  |  |  |
| Female | 16,521 (49.8%) | 3,123 (49.6%) | 4,122 (49.6%) | 4,960 (49.7%) | 2,844 (49.5%) | 1,472 (52.0%) |
| Male | 16,619 (50.1%) | 3,165 (50.3%) | 4,181 (50.3%) | 5,022 (50.3%) | 2,892 (50.4%) | 1,359 (48.0%) |
| Other/Unknown | 19 (0.1%) | 5 (0.1%) | 4 (0.0%) | 4 (0.0%) | 5 (0.1%) | 1 (0.0%) |
| County |  |  |  |  |  |  |
| Brazoria | 2,801 (8.4%) | 600 (9.5%) | 618 (7.4%) | 923 (9.2%) | 439 (7.6%) | 221 (7.8%) |
| Chambers | 256 (0.8%) | 66 (1.0%) | 84 (1.0%) | 61 (0.6%) | 32 (0.6%) | 13 (0.5%) |
| Fort Bend | 3,187 (9.6%) | 463 (7.4%) | 752 (9.1%) | 994 (10.0%) | 660 (11.5%) | 318 (11.2%) |
| Galveston | 4,165 (12.6%) | 1,049 (16.7%) | 1,137 (13.7%) | 1,122 (11.2%) | 573 (10.0%) | 284 (10.0%) |
| Harris | 18,493 (55.8%) | 3,410 (54.2%) | 4,759 (57.3%) | 5,631 (56.4%) | 3,247 (56.6%) | 1,446 (51.1%) |
| Liberty | 719 (2.2%) | 157 (2.5%) | 213 (2.6%) | 208 (2.1%) | 94 (1.6%) | 47 (1.7%) |
| Montgomery | 3,039 (9.2%) | 523 (8.3%) | 697 (8.4%) | 921 (9.2%) | 635 (11.1%) | 263 (9.3%) |
| Waller | 271 (0.8%) | 25 (0.4%) | 46 (0.6%) | 125 (1.3%) | 54 (0.9%) | 21 (0.7%) |
| Unknown | 228 (0.7%) | 0 (0.0%) | 1 (0.0%) | 1 (0.0%) | 7 (0.1%) | 219 (7.7%) |
| Diagnostic encounter type* |  |  |  |  |  |  |
| Emergency | 15,435 (46.5%) | 3,135 (49.8%) | 3,660 (44.1%) | 3,877 (38.8%) | 3,361 (58.5%) | 1,402 (49.5%) |
| Inpatient | 3,565 (10.8%) | 541 (8.6%) | 821 (9.9%) | 659 (6.6%) | 1,032 (18.0%) | 512 (18.1%) |
| Outpatient | 10,555 (31.8%) | 1,158 (18.4%) | 1,566 (18.9%) | 2,141 (21.4%) | 3,833 (66.8%) | 1,857 (65.6%) |
| **RSV Laboratory Tests Resulted** | **N=165,456** | **N=44,902** | **N=54,014** | **N=43,551** | **N=16,488** | **N=6,501** |
| Total RSV Antigen Tests Resulted | 65,115 (39.4%) | 12,550 (27.9%) | 21,664 (40.1%) | 18,424 (42.3%) | 9,097 (55.2%) | 3,380 (52.0%) |
| Positive RSV Antigen Tests, n (% of total antigen tests) | 9,009 (13.8%) | 1,917 (15.3%) | 3,650 (16.8%) | 2,342 (12.7%) | 782 (8.6%) | 318 (9.4%) |
| Total RSV PCR Tests Resulted | 100,341 (60.6%) | 32,352 (72.1%) | 32,350 (59.9%) | 25,127 (57.7%) | 7,391 (44.8%) | 3,121 (48.0%) |
| Positive RSV PCR Tests, n (% of total PCR tests) | 2,508 (1.5%) | 660 (1.5%) | 848 (1.6%) | 641 (1.5%) | 243 (1.5%) | 116 (1.8%) |
| **Patients with ≥ 1 RSV antigen or PCR test resulted** | **N=104,662** | **N=29,680** | **N=30,409** | **N=27,330** | **N=11,998** | **N=5,245** |
| Patients with ≥ 1 RSV antigen test resulted, n (% of total patients with a test) | 48,680 (46.5%) | 9,652 (32.5%) | 14,471 (47.6%) | 13,961 (51.1%) | 7,487 (62.4%) | 3,109 (59.3%) |
| Patients with ≥ 1 positive RSV antigen test, n (% of total patients with an antigen test) | 7,648 (15.7%) | 1,625 (16.8%) | 2,712 (18.7%) | 2,229 (16.0%) | 770 (10.3%) | 312 (10.0%) |
| Patients with ≥ 1 RSV PCR test resulted, n (% of total patients with a test) | 57,834 (55.3%) | 20,382 (68.7%) | 16,640 (54.7%) | 13,946 (51.0%) | 4,661 (38.8%) | 2,205 (42.0%) |
| Patients with ≥ 1 positive RSV PCR test, n (% of total patients with a PCR test) | 1,946 (3.4%) | 514 (2.5%) | 583 (3.5%) | 515 (3.7%) | 221 (4.7%) | 113 (5.1%) |
| Patients with ≥ 1 positive RSV test, n (% of total patients with a test) | 9,544 (9.1%) | 2,131 (7.2%) | 3,270 (10.8%) | 2,730 (10.0%) | 989 (8.2%) | 424 (8.1%) |

Case definitions: RSV episodes defined per person per week as any qualifying RSV diagnosis (ICD-10 or SNOMED codes) recorded more than 2 weeks from the start of the previous episode. Each patient can contribute only one RSV episode within a 3-week period. Laboratory tests include all diagnostic antigen or PCR-based laboratory tests resulted. Inconclusive, failed, or unknown results were excluded. Note that tests for both RSV A and B are included, which may inflate tests per person per encounter. Study population: Individuals with a self-reported residential address within Harris, Fort Bend, Galveston, Montgomery, Liberty, Waller, Chambers, and Brazoria counties and at least one target, either diagnosis or laboratory test report, recorded in the HIE between January 1, 2022, and August 26, 2026.
\*Diagnostic encounter type defined non-exclusively. For example, 'Inpatient COVID-19' includes all patients with at least one COVID-19 diagnosis recorded at an inpatient encounter.

#### Influenza

Influenza case counts were relatively consistent over the study period, with the exception of 2025, which was approximately twice as high as the other years (Table 3). Unusually, the proportion of patients with at least one influenza-associated outpatient encounter showed a large increase between 2024 and 2025. Around 80% of the 1,318,235 Influenza A and B tests resulted in our study population were antigen tests. Over the years, influenza cases peaked between October and February, with marked variability in timing of epidemic peaks (Figure 2). Test positivity rates for Influenza A and B exhibited separate trends, with Influenza A peaking every year and Influenza B displaying a strong peak in 2023 and moderate peaks in late spring of 2025 and early spring of 2026. Case count trends largely aligned temporally with peaks in both Influenza A and B, though the magnitude of cases did not appear to correlate strongly with the percent positivity (Figure 3). Influenza-associated hospitalizations were highest among older adults. Hospitalizations associated with influenza were highest in January 2025.

**Table 3:**
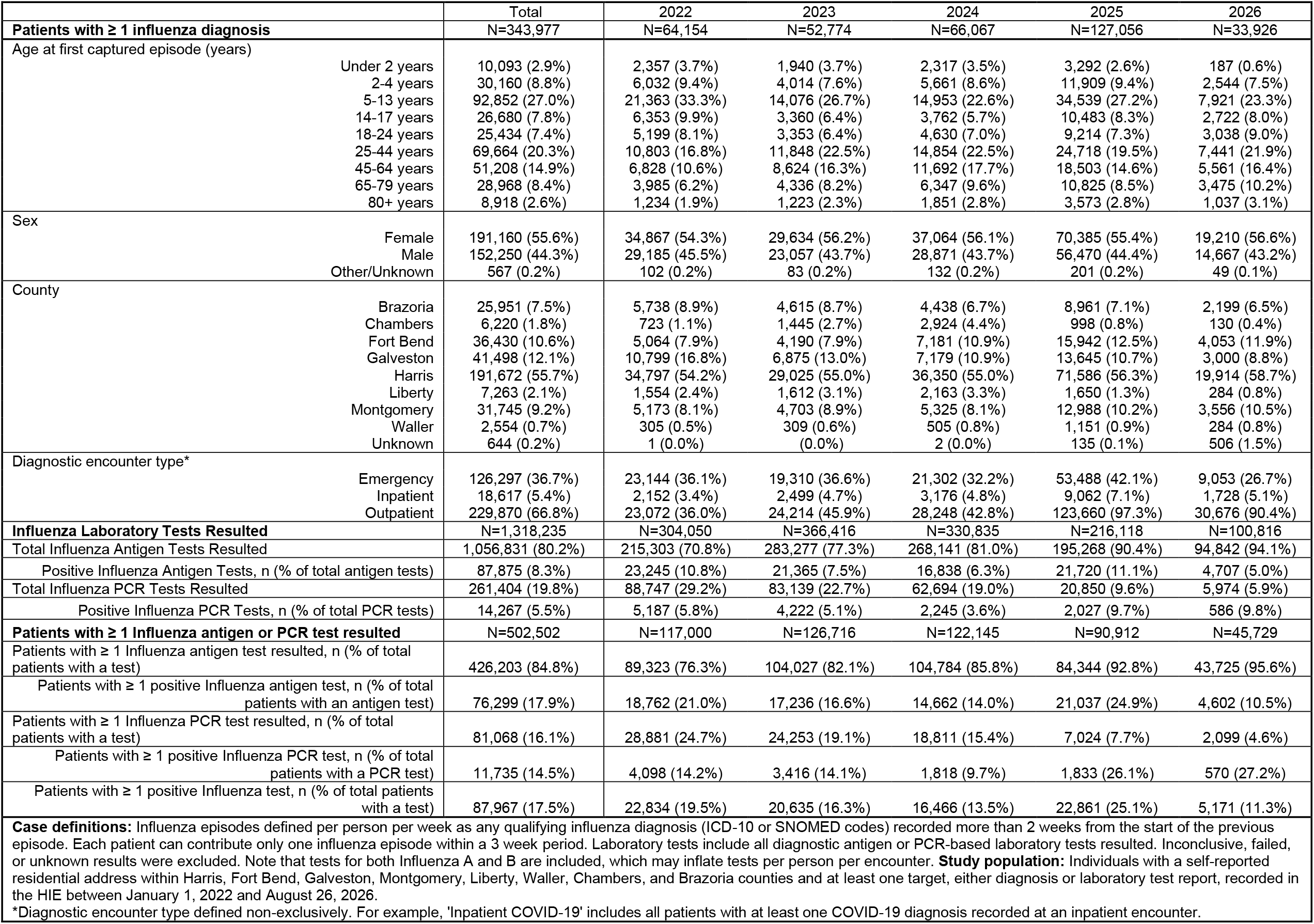
Influenza episodes captured in HIE-based surveillance platform, January 2022 to August 2026.

## Discussion

In this study, we describe the development, validation, and implementation of a flexible, scalable, near-real-time EHR-based surveillance platform. By leveraging a large regional network of healthcare organizations, we effectively captured a representative proportion of clinically attended COVID-19, influenza, and RSV episodes in the Greater Houston area. These results demonstrate the capacity of health information exchange networks to execute system-agnostic queries utilizing health data interoperability standards and robust pipelines to extract, process, and analyze complex EHR data quickly and efficiently. The HIE infrastructure overcomes the siloing of data within health systems, capturing patients’ movement across care settings to more accurately measure public health surveillance metrics.

Our platform was able to not only characterize trends in incidence, laboratory test positivity, and hospitalizations associated with COVID-19, RSV, and influenza across the Greater Houston Area, but also identify important subpopulations relevant to infectious disease prevention and control. The data show expected seasonal trends in these respiratory infections, as well as unforeseen events, including the COVID-19 late winter peaks fading and summer peaks shifting later into autumn over the 2022-2026 study period. The RSV case trends additionally show an unusual summer peak in 2022, which corresponds to the lifting of pandemic-era restrictions earlier that spring. Perhaps most importantly, hospitalizations associated with RSV appear to have decreased dramatically among infants in the 2025/2026 season, both compared to previous years, and proportionally to other age groups. This trend must be further explored in the context of the novel RSV vaccine and monoclonal antibody campaigns which launched in the summer of 2023, targeting infants under 8 months, pregnant women between 32 and 36 weeks gestation, and older adults [21, 22].

The major challenge in designing and interpreting EHR-based surveillance programs is the lack of a clear ‘denominator’; the source population for an EHR dataset is often poorly defined, and healthcare utilization data by its nature is biased towards more severe illness [3]. These limitations can be somewhat mitigated when surveillance is conducted within a mature HIE infrastructure, if the baseline population can be established before the outbreak or event and biases can be incorporated into the interpretation of findings [23]. By pairing this novel surveillance platform with rigorous assessment of the source population covered within a regional HIE, we can more confidently interpret case counts, laboratory test trends, and hospitalization rates across multiple demographic metrics. Future investigations will extend this premise by calculating sampling weights to enable population-based adjustment of case data.

No surveillance dataset contains complete information on disease burden in a population. We demonstrated that even for targets with lower overall reporting proportions, such as laboratory test data, our platform was able to detect interpretable, community-level trends in line with traditional public health reporting for the same pathogens [24]. Our success likely stems from both the overall volume of patient data within the network as well as the hierarchical reporting structure. Specifically, data streams such as laboratory test results are typically available or missing at the healthcare institution level, not the individual patient level, mitigating the potential selection bias even in an incomplete sampling frame.

In conclusion, our findings demonstrate that health information exchange can facilitate enhanced population-level surveillance for infectious diseases, despite heterogenous reporting across health systems. The results from this surveillance platform demonstrate the great potential of cross-system data exchange for advancing population health. By breaking down institutional and system-level data silos, platforms such as ours can equip public health authorities and health networks with real-time, actionable epidemiological intelligence, providing a vital blueprint for resilient regional health security and disease prevention.

## Data Availability

The datasets presented in this article are not readily available because clinical data cannot be shared publicly because of patient confidentiality concerns as imposed by the University of Texas Health Science Center at Houston Committee for the Protection of Human Subjects. Requests to access de-identified data can be made to which will be evaluated on a case by case basis in line with institutional policies.

## Notes

### Competing Interest Statement

The authors have declared no competing interest.

### Funding

- S.B. 1780, 87th Legislature, 2021 Reg. Sess. (Texas 2021)

## References

1. Sarwar T, Seifollahi S, Chan J, Zhang X, Aksakalli V, Hudson I, Verspoor K, Cavedon L. The secondary use of electronic health records for data mining: data characteristics and challenges. ACM Computing Surveys (CSUR). 2022 Jan 18;55(2):1–40.

2. Sauer CM, Chen LC, Hyland SL, Girbes A, Elbers P, Celi LA. Leveraging electronic health records for data science: common pitfalls and how to avoid them. The Lancet Digital Health. 2022 Dec 1;4(12):e893–8.

3. Casey JA, Schwartz BS, Stewart WF, Adler NE. Using electronic health records for population health research: a review of methods and applications. Annual review of public health. 2016 Mar 18;37(1):61–81.

4. Lin CT, Bookman K, Sieja A, Markley K, Altman RL, Sippel J, Perica K, Reece L, Davis C, Horowitz E, Pisney L. Clinical informatics accelerates health system adaptation to the COVID-19 pandemic: examples from Colorado. Journal of the American Medical Informatics Association. 2020 Dec;27(12):1955–63.

5. Hirsch AG, Conderino S, Crume TL, Liese AD, Bellatorre A, Bendik S, Divers J, Anthopolos R, Dixon BE, Guo Y, Imperatore G. Using electronic health records to enhance surveillance of diabetes in children, adolescents and young adults: a study protocol for the DiCAYA Network. BMJ open. 2024 Jan;14(1):e073791.

6. Comer KF, Gibson PJ, Zou J, Rosenman M, Dixon BE. Electronic health record (EHR)-based community health measures: an exploratory assessment of perceived usefulness by local health departments. BMC Public Health. 2018 May 22;18(1):647.

7. Shapiro MJ, Kannry MJ, Lipton MM, Goldberg ME, Conocenti MP, Stuard MS, Wyatt JB. Approaches to patient health information exchange and their impact on emergency medicine. Annals of emergency medicine. 2006 Oct 1;48(4):426–32.

8. Shapiro JS, Mostashari F, Hripcsak G, Soulakis N, Kuperman G. Using health information exchange to improve public health. American journal of public health. 2011 Apr;101(4):616–23.

9. Poirot E, Mills CW, Fair AD, Graham KA, Martinez E, Schreibstein L, Talati A, McVeigh KH. Evaluation of a health information exchange system for microcephaly case-finding—New York City, 2013—2015. Plos one. 2020 Aug 17;15(8):e0237392.

10. Khurshid A, Kendrick D, Pillai HS, Sun J, Oliveira E, Jaquish A, Dutcher SK, Bright P, Wang Y, Li J, Austin T. Health Information Exchanges (HIEs) as Novel Sources for Population-Based Post Marketing Surveillance of Medical Products: A Pilot Study from the FDA Sentinel Innovation Center: A. Khurshid et al. Drug Safety. 2026 Apr 13:1–1.

11. Vest JR, Ben-Assuli O. Prediction of emergency department revisits using area-level social determinants of health measures and health information exchange information. International journal of medical informatics. 2019 Sep 1;129:205–10.

12. Garg N, Kuperman G, Onyile A, Lowry T, Genes N, DiMaggio C, Richardson L, Husk G, Shapiro JS. Validating health information exchange (HIE) data for quality measurement across four hospitals. InAMIA Annual Symposium Proceedings 2014 Nov 14 (Vol. 2014, p. 573).

13. Sandoval MN, Mikhail JL, Fink MK, Tortolero GA, Cao T, Ramphul R, Husain J, Boerwinkle E. Social determinants of health predict readmission following COVID-19 hospitalization: a health information exchange-based retrospective cohort study. Frontiers in Public Health. 2024 Mar 27;12:1352240.

14. Sandoval MN, Mikhail JL, Fink MK, Tortolero GA, Cao T, Ramphul R, Husain J, Boerwinkle E. Social determinants of health predict readmission following COVID-19 hospitalization: a health information exchange-based retrospective cohort study. Frontiers in Public Health. 2024 Mar 27;12:1352240.

15. U.S. Census Bureau. “Profile of General Population and Housing Characteristics: 2020 Census Demographic Profile (Table DP1).” 2020 Decennial Census, 2023, https://data.census.gov/table/DECENNIALDP2020.DP1.

16. U.S. Census Bureau. “2019-2023 American Community Survey 5-Year Estimates: Table B01003 Total Population.” Texas.

17. Li W, Zhang M, Cheng J, Jackson SL, Vaughan AS, Klompas M, Carton TW, Nauman E, Mendoza L, Adhikari A, Donovan J. Weighted EHR-based prevalence estimates for hypertension at the state and local levels in Louisiana. BMC Public Health. 2025 Feb 3;25(1):432.

18. U.S. Census Bureau. 2020. Understanding and Using American Community Survey Data: What Researchers Need to Know. Washington, DC: U.S. Census Bureau

19. Centers for Disease Control and Prevention (U.S.). MMWR Week Log. Atlanta, GA: US Department of Health and Human Services, CDC.

20. Walker K, Herman M (2026). tidycensus: Load US Census Boundary and Attribute Data as ‘tidyverse’ and ‘sf’-Ready Data Frames. R package version 1.8.0, https://walker-data.com/tidycensus/

21. Venkatesan P. First RSV vaccine approvals. The Lancet Microbe. 2023 Aug 1;4(8):e577.

22. Payne AB, Watts JA, Mitchell PK, Dascomb K, Irving SA, Klein NP, Grannis SJ, Ong TC, Ball SW, DeSilva MB, Natarajan K. Respiratory syncytial virus (RSV) vaccine effectiveness against RSV-associated hospitalisations and emergency department encounters among adults aged 60 years and older in the USA, October, 2023, to March, 2024: a test-negative design analysis. The Lancet. 2024 Oct 19;404(10462):1547–59.

23. Dixon BE, Wools-Kaloustian KK, Fadel WF, Duszynski TJ, Yiannoutsos C, Halverson PK, Menachemi N. Symptoms and symptom clusters associated with SARS-CoV-2 infection in community-based populations: Results from a statewide epidemiological study. PLoS One. 2021 Mar 24;16(3):e0241875.

24. Centers for Disease Control and Prevention. (n.d.). Coronavirus Disease 2019 (COVID-19) hospitalization surveillance network (COVID-NET). U.S. Department of Health and Human Services.

